# Persistent alveolar type 2 dysfunction and lung structural derangement in post-acute COVID-19

**DOI:** 10.1101/2022.11.28.22282811

**Authors:** André F. Rendeiro, Hiranmayi Ravichandran, Junbum Kim, Alain C. Borczuk, Olivier Elemento, Robert E. Schwartz

**Author notes:** Co-first authors (André F. Rendeiro, Hiranmayi Ravichandran). Co-senior authors (Alain C. Borczuk, Olivier Elemento, Robert E. Schwartz).

## Abstract

SARS-CoV-2 infection can manifest as a wide range of respiratory and systemic symptoms well after the acute phase of infection in over 50% of patients. Key questions remain on the long-term effects of infection on tissue pathology in recovered COVID-19 patients. To address these questions we performed multiplexed imaging of post-mortem lung tissue from 12 individuals who died post-acute COVID-19 (PC) and compare them to lung tissue from patients who died during the acute phase of COVID-19, or patients who died with idiopathic pulmonary fibrosis (IPF), and otherwise healthy lung tissue. We find evidence of viral presence in the lung up to 359 days after the acute phase of disease, including in patients with negative nasopharyngeal swab tests. The lung of PC patients are characterized by the accumulation of senescent alveolar type 2 cells, fibrosis with hypervascularization of peribronchial areas and alveolar septa, as the most pronounced pathophysiological features. At the cellular level, lung disease of PC patients, while distinct, shares pathological features with the chronic pulmonary disease of IPF. which may help rationalize interventions for PC patients. Altogether, this study provides an important foundation for the understanding of the long-term effects of SARS-CoV-2 pulmonary infection at the microanatomical, cellular, and molecular level.

## Main text

Coronavirus disease 2019 (COVID-19) is now recognized as a systemic disease affecting the lung and several extrapulmonary organs^1,2^. Its effects can be prolonged after acute infection developing into a syndrome of post-acute sequelae of COVID-19 (PASC)^3–6^. The cumulative frequency of PASC symptoms such as fatigue, cough, joint and chest pain, and dyspnea from patients that survive acute COVID-19^7–10^ is a considerable burden in loss of health and quality of life and impacts upwards of 50% of patients following infection^11,12^. Recent studies of health records have tallied the cumulative burden of PASC symptoms, revealing increased risk for developing comorbidities systemically, in a manner dependent on the severity of the acute phase of disease^11^. Blood profiling of PASC patients revealed lasting immune, serological and hormonal differences in comparison with convalescent COVID-19 patients without PASC^13^. However, the pathophysiological roots of the pervasiveness of dyspnea and other symptoms that manifest at the respiratory level in individuals suffering from post-acute COVID-19 is not well understood. We had previously observed increased fibrosis^14^ as well as widespread infection and injury of alveolar type 2 (AT-2) cells in the lungs of severe COVID-19 patients^15^. We now hypothesize that long-term sequelae of SARS-CoV-2 infection may share similarities with chronic lung disease such as usual interstitial pneumonia associated with idiopathic pulmonary fibrosis (UIP/IPF). Indeed, accumulating evidence suggests a role of AT-2 dysfunction driving fibrosis in UIP/IPF^16,17^. To test this hypothesis, we used multiplexed imaging to conduct a multiscale comparative analysis of post-mortem lung samples of patients of acute COVID-19, patients that died after the acute phase of COVID-19, as well as post-mortem samples of IPF patients, thus seeking to uncover potentially shared pathophysiological features from the microanatomical to the single-cell level.

### Persistent SARS-CoV-2 presence associated with AT-2 cell senescence

To shed light into the process of lung recovery after COVID-19, we investigated a cohort of 12 patients with respiratory symptoms who died incidentally after an acute period of COVID-19 (post-COVID, PC) (**Figure 1a, Supplementary Table 1**). We divided the PC group based on the last nasopharyngeal (NP) test before the time of death as positive (PC-pos) or negative (PC-neg). The PC-neg group of patients had a clinical course with a mean of 77 days, and a mean of 4 negative COVID-19 NP tests before death (range 2-8) (**Figure 1b**). The PC-pos group had a clinical course with a mean of 314 days, with an acute infection period in early 2020 (**Figure 1b**). As reference groups, we included samples from individuals who died without lung disease (n = 2), patients with UIP/IPF (n = 2), and patients who died during the acute phase of COVID-19 (n = 4) (**Figure 1a**). From this latter group, we included patients who died after 1 and 5 days (early COVID-19), and 35 and 36 days (late COVID-19) after symptom onset (**Figure 1a**).

**Figure 1:**
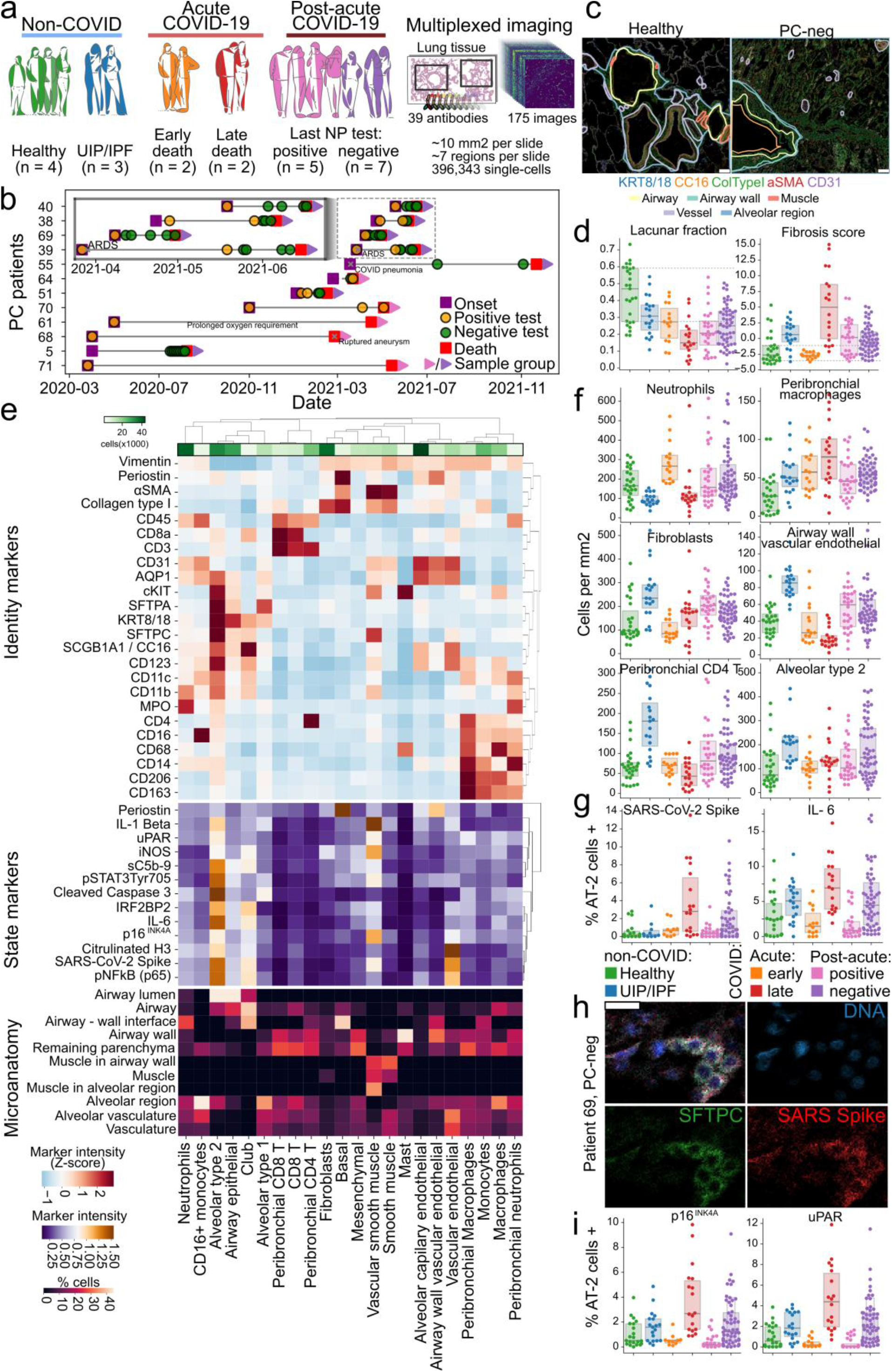
Long-term presence of SARS-CoV-2 in the lungs of post-acute COVID-19 patients. **a)** Conceptual description of the cohort of patients in the study and process of acquisition of imaging mass cytometry data. **b**) Timelines of PC patients including major events during the courses of disease. **c)** Representative images of lung tissue imaged with imaging mass cytometry for a healthy lung sample (left), and PC-neg with outline of microanatomical domains overlaid. Scale bar represents 100 μm. **d)** Quantification of the lacunar fraction (left), and fibrosis score (right) for each image grouped by sample group. **e)** Clustered heatmap of cell types identified across all images in the IMC dataset. The upper part contains markers associated with cell type identity, the middle with cellular state, and the lower part of the heatmap represents the relative enrichment of each cell type in the delineated microanatomical domains of the lung. **f)** Absolute abundance (cell density per square millimeter) of selected cell types across sample groups. **g)** Percentage of AT-2 cells from different sample groups which are positive for SARS-CoV-2 Spike protein (left), or IL-6 (right). **h)** Illustration of infected AT-2 cells in the lung of a patient from the PC-neg group. The upper left image represents the merge of all remaining three. Scale bar represents 25 μm. **i)** Percentage of AT-2 cells from different sample groups which are positive for p16 (left), or uPAR (right).

To investigate the cellular and microanatomical pathophysiology of lung tissue from the patients, we designed a panel of 39 antibodies for imaging mass cytometry^18^ (**Supplementary Table 2**). We then generated 175 multiplexed images with 1 μm resolution, comprising 221 mm^2^ of tissue, from which 396,343 single cells could be identified with high-confidence (**Figure 1a, Supplementary Figure 1**). To augment the cells in our images with structural context of the tissue, we delineated boundaries for microanatomical domains such as airways, vessels, muscle, and alveolar region in the multiplexed images (**Figure 1c, Supplementary Figure 2**). At the microanatomical level, we noticed significantly less space dedicated to gas exchange and blood circulation for all pathological sample groups including PC patients when compared with healthy lung (**Figure 1c-d, Supplementary Table 3**). We also noticed increased fibrosis (with the exception of early acute COVID-19), which we had previously reported^14^. Surprisingly, the level of microanatomical pathology in PC patients was comparable to both early and late groups of acute COVID-19 and UIP/IPF (**Figure 1c-d**).

To better understand the cellular changes in PC patients, we leveraged the similarity of marker expression between single-cells to establish 43 cell clusters which we further categorized into 22 cell types depending on marker intensity and microanatomical context (**Figure 1e, Supplementary Figure 3**). The inclusion of markers specific to various types of epithelial cells in the lung such as SFTPA, SFTPC, and SCGB1A1, together with microanatomical annotation allowed us to resolve cellular identity to a higher degree than previously possible^14^, particularly for epithelial and endothelial cell types. We observed both COVID-induced changes for several cell types, as well as commonalities between acute COVID-19, post-COVID-19 or UIP/IPF (**Supplementary Figure 4**). For instance, neutrophils - which are a hallmark in lungs of early acute COVID-19 patients^14,19,20^ - are significantly higher in PC patients in comparison with late acute COVID-19 patients. Conversely, predominantly peribronchial, interstitial macrophages - which are the hallmark of late acute COVID-19^14,15^ - are significantly increased in all disease groups in comparison with healthy lung (**Figure 1f, Supplementary Figure 4**). Similar to the UIP/IPF group, we also observed increased density of fibroblasts in PC in comparison with healthy lung, as well as higher density of vascular endothelial cells predominantly in the airway walls and CD4+ T-cells. The latter two cell types were increased in PC not only in comparison to healthy lung, but also to acute COVID-19 and regardless of the status of last NP test of the patient (**Figure 1f, Supplementary Figure 4**). These observations suggest persistent changes in both the immune and structural compartments of the lung long after acute infection and associated with long-term post-acute COVID-19 sequelae.

In some samples we also observed a higher density of alveolar type 2 (AT-2) cells in PC, particularly in the PC-neg group (**Figure 1f, Supplementary Figure 4, Supplementary Table 3**). Since AT-2 cells are the cell type most infected by SARS-CoV-2 in the lung^14,15^, and their injury leads to accumulation of dysfunctional cells^15^, we checked whether AT-2 cells of PC samples had detectable virus. Immunoreactivity for SARS-CoV-2 Spike protein was found most predominantly in surfactant C^+^ (SFTPC) AT-2 cells of samples from the PC-neg group (**Figure 1g-h, Supplementary Figure 5, Supplementary Figure 6a-b**), to a degree comparable to late COVID-19. Furthermore, in AT-2 cells predominantly of late COVID-19 and PC-neg, we also detected the highest levels of the inflammatory marker IL-6 (**Figure 1g**), which is upregulated in SARS-CoV-2 infected cells^14^.

SARS-CoV-2 infection is responsible for the injury and accumulation of dysfunctional AT-2 cells^15^, which are responsible to maintain and regenerate the alveolar niche of the lung^21–24^. Accumulating evidence points to the depletion or senescence of AT-2 cells in IPF as responsible for the fibrotic phenotype of the pathology^16,17^. We investigated whether AT-2 cells were bearing markers of cellular senescence. Indeed, the senescence-associated p16^INK4A^ ^25–27^ and urokinase-type plasminogen activator receptor (uPAR)^28,29^ were significantly increased in AT-2 cells from late COVID-19, and PC-neg patients in a pattern matching the rate of SARS-CoV-2 infection in each group (**Figure 1i, Supplementary Table 3**). Furthermore, we also observed significant increases of the senescence markers for these patient groups in fibroblasts, mesenchymal and vascular endothelial cells (**Supplementary Figure 5b**). Finally, we also observed ectopic AT-2 cells in the lumen of airways and alveoli (**Figure 1e**). Such anomalous structural derangement has been observed in chronic lung disease as well^30,31^.

To orthogonally validate the presence of SARS-CoV-2 Spike protein, we performed *in situ* hybridization, immunohistochemistry, and PCR of formalin-fixed paraffin embedded lung tissue for SARS-CoV-2. We observed that cell types with the highest signal in the imaging data also had the largest difference between samples labeled as positive by the orthogonal methods, showing positive, significant agreement between multiplexed imaging and standard methods of tissue analysis (**Supplementary Figure 6c**). The agreement between orthogonal assays reinforces the validity of our observations of specific and persistent changes in microanatomy, cellular composition, and expression in the lungs of individuals who died post-acute COVID-19.

### Derangement of vascular and epithelial lung structure in PC patients

We observed similarity in specific aspects of lung organization and composition between acute COVID-19 and patients who died post-COVID-19 (increased neutrophils and macrophages), as well between UIP/IPF and PC (increased fibroblasts and AT-2 injury) (**Figure 1d-g,i**). We sought to more systematically compare the disease entities by observing the global similarity between samples (**Figure 2a-b, Supplementary Figure 7**), and disease groups (**Figure 2c**). UIP/IPF and late COVID-19 were outstanding groups, whereas healthy lung, acute COVID-19 and PC were more similar to each other, with more heterogeneous groups (**Figure 2a-b**). We nonetheless observed that PC individuals whose last NP test was positive (PC-pos) were most globally similar to individuals with UIP/IPF, whereas the PC-neg group was more globally similar to the early COVID-19 group. This could be due to persistent active viral-driven disease or differences in time since index infection between the groups, which could have different levels of fibrosis.

**Figure 2:**
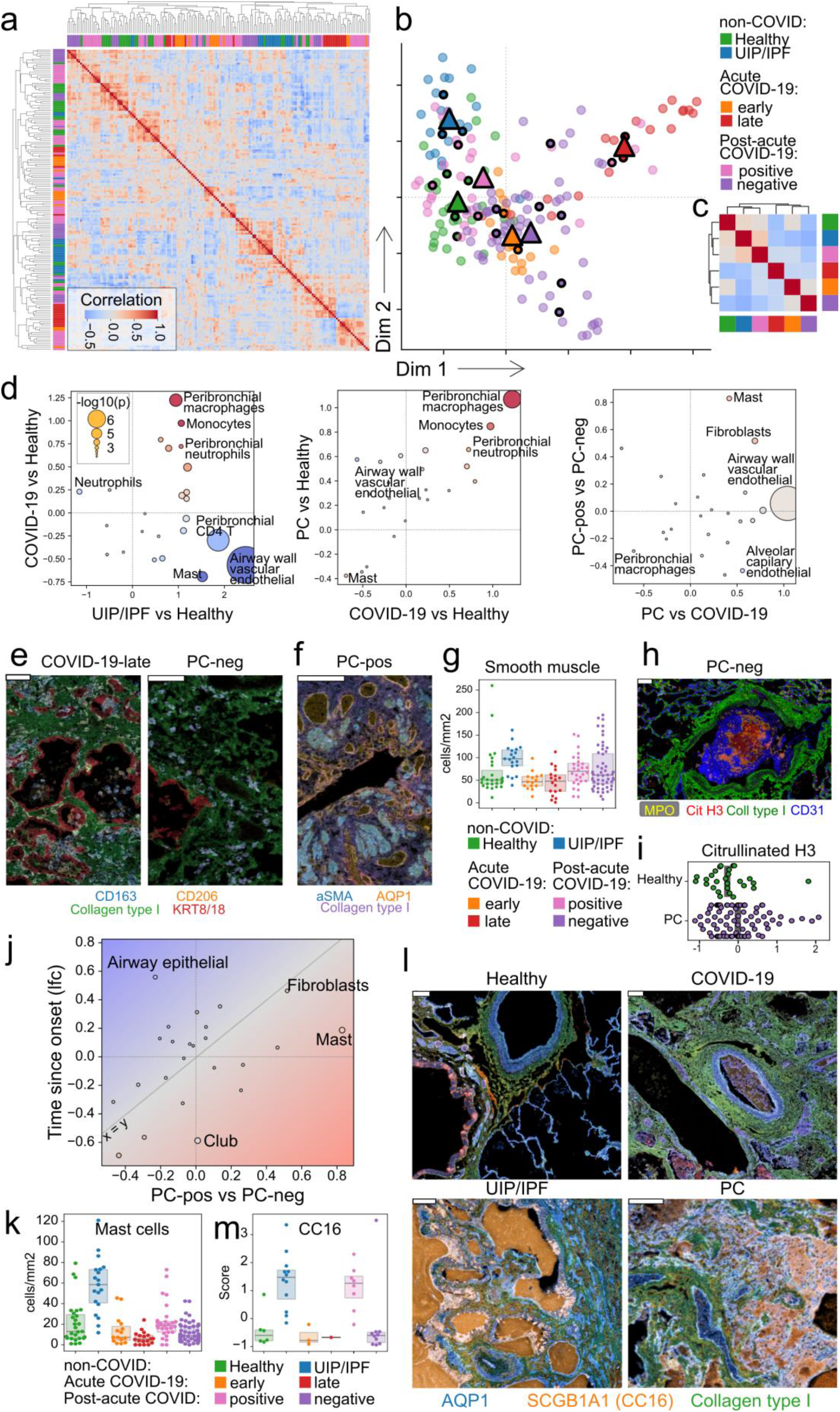
Comparative microanatomical analysis of lung identifies lasting structural derangement. **a**) Clustered heatmap of pairwise correlation between images based on the abundance of cell types. Image group identity is marked by colors in rows and columns with legend in b). **b**) Two-dimensional projection of the same data as in a) using Spectral Embedding. **c**) Same as a) but grouped by disease group. **d**) Scatter plot of effect sizes estimated from comparing various disease groups. Point size is related to the significance of change, while color displays agreement between comparisons (agreement in red, disagreement in blue tones). **e**) Representative images of macrophages in acute COVID and PASC. **f**) Image highlighting vascular network and muscularization of lung tissue in PASC sample. **g**) Absolute abundance of smooth muscle cells in lung tissue from various disease groups. **h**) Exemplary image of blood clot where MPO^+^ neutrophils can be seen in the lumen with Citrullinated H3 from NETs in proximity. **i**) Quantification of extracellular citrullinated H3 signal in healthy lung and PASC. **j**) Scatter plot of estimated effect sizes of cell type abundance changes compared between patients in the PC-pos and -neg groups (x-axis), and within PC patients along the time since disease onset (y-axis). **k**) Absolute abundance of mast cells in the lungs of patients from the various disease groups. **l**) Representative images of secreted CC16 for various disease groups. AQP1 and Collagen shown as reference. **m**) Amount of extracellular CC16 (SCGB1A1) across disease groups for images containing airways.

To shed light into the features underlying the differences and similarities between disease groups, we first contrasted the cellular composition of the samples between them (**Figure 2d**). As previously described, we found increased density of peribronchial macrophages and monocytes in acute COVID-19 in comparison with healthy lung^14^, while vascular remodeling^32^, CD4+ T-cells^33^ and mast cells^34^ are hallmarks of IPF (**Figure 2d**, left). We found overall agreement between changes in cell composition of post-acute and acute COVID-19 with CD163^+^/CD206^+^ myeloid cells dominating changes for both disease groups in comparison to healthy lung (**Figure 2d**, center, **Figure 2e**). However, we found that vascular endothelial cells predominantly in airway walls were increased in density only in PC (**Figure 2d**, center), suggesting that as in IPF, vascular remodeling may be of pathophysiological relevance. Indeed, we observed not only widespread ectopic appearance of microvasculature but also significantly more smooth muscle cells in peribronchial regions and alveolar septa (**Figure 2f-g**). The appearance of both ectopic AT2-cells (**Figure 1d**) and microvasculature highlights the increased derangement of lung structure in PC. Furthermore, since neutrophil deployment of extracellular traps (NET) is known to drive vascular inflammation^35,36^ and neutrophils in PC remain increased in comparison with healthy lungs, we sought to quantify the amount of NETs in the lung. We observed that the total amount of extracellular citrullinated H3, a marker of neutrophil NETs^37^ was significantly higher in PC when compared with healthy lung (**Figure 2h-i**). These results highlight the degree of long-term cellular and structural derangements in post-acute COVID-19 patients.

Lung healing through regeneration or repair is a complex process unfolding over the timescale of months^38^. It is therefore plausible that patients who died during the acute phase of COVID-19 compared with post-acute patients have distinct features due to the continuous healing process, in addition to potential specific processes related with either acute or post-acute COVID-19. We sought to deconvolute time- and pathology-specific processes in our multiplexed imaging data by focusing only on the group of post-acute COVID-19 patients and contrasting changes associated with the time since the acute phase of disease across all patients, and differences between the two PC subgroups (**Figure 2j**). This analysis revealed an overall agreement between the cellular changes associated with the two PC groups and the time since disease onset, with increases in fibroblasts over time consistent with the fibrotic-mediated scarring during lung repair. However, we also detected changes in cell abundance between PC groups which were independent of the time since symptom onset. Mast cells were more abundant in the PC-pos group than PC-neg independently of the time, in some cases to a similar degree as in UIP/IPF (**Figure 2k**). We also detected changes in the airway epithelium which were mostly driven by time rather than differences between PC groups. However, a common feature of IPF is the ectopic appearance of airway epithelial cells (bronchiolization), and their secretion of CC16/SCGB1A1 particularly in Club cells^39–42^. We had observed that similarly to IPF, some images from PC patients showed extensive secreted CC16 in peribronchial regions (**Figure 2l**). We therefore quantified the amount of secreted SCGB1A1 and found levels in the PC-pos group comparable to IPF and unlike that seen in healthy lung or acute COVID-19 (**Figure 2m**). These observations highlight a globally aberrant microanatomical and cellular state of lungs of post-acute COVID-19 patients and identifies similarities with pathological features of UIP/IPF particularly for patients with prolonged active viral disease.

## Discussion

Our analysis of post-mortem lung tissue from the microanatomical to single-cell level with multiplexed imaging provides a thorough examination of acute infection and host response, as well as the chronic pathological processes that manifest at long-term after acute infection.

In post-acute COVID-19 (PC) patients followed for a period of up to 399 days, we identified surprisingly persistent loss of lung lacunarity (as a proxy for pulmonary capacity), fibrosis, and perturbed immune and structural cellular components of the lung. Of particular note, we identified persistent presence of SARS-CoV-2 epitopes in the lung of PC patients, which did not necessarily agree with the status of the last nasopharyngeal test of the patients. It however agrees with recent reports showing viral factors present in circulation^43^ and in the gut^44^ of PASC patients. We also observed that SARS-CoV-2 presence is linked with the activation of senescence in AT-2, mesenchymal, endothelial, and fibroblast cells as detected by the upregulation of p16^INK4A^, uPAR, and IL-6 which is part of the senescence associated secretory profile (SASP). SARS-CoV-2 induction of senescence in AT-2 cells had been suggested in acute disease^45–47^, but its long-term persistence after the acute disease raises the question of whether AT-2 senescence provides a reservoir for SARS-CoV-2 -something that would require future validation using viral replication assays. Furthermore, the fact that mesenchymal cells in addition to AT-2s display increased senescence suggests a “double-hit” for the maintenance of epithelial cells in the lung since they can promote AT-2 self-renewal^48,49^ and relieve AT-2 senescence^50^.

Overall, we observed that PC cases showed distinct features in comparison to healthy lung or acute COVID-19, with extensive derangement of the vascular network characterized by increased microvascularization and NET-induced vascular damage. Thrombosis as well as intussusceptive and sprouting angiogenesis have been found in acute COVID-19^51^ at higher levels than in Influenza patients. The fact that in our data we find the level of vascular derangement higher in post-acute than in acute COVID-19 suggests that vascular remodeling is a continuously acting process during post-acute COVID disease.

While UIP/IPF and sequelae of SARS-CoV-2 infection develop at largely different time scales (years and months respectively), we identified several common aspects between the diseases such as increased fibrosis and fibroblast abundance, vascular remodeling, increase of Mast cells, and upregulation of CC16/SCGB1A1 secretion. Mast cells play a pivotal role in IPF, with particular importance to the establishment of the fibrotic phenotype^52,53^. Mast cell involvement in PASC has been suggested based on comparative analysis of symptomatic questionnaires^54^ and molecular analysis of circulatory factors^55^. The secreted Club cell marker CC16/SCGB1A1 is overexpressed in IPF^42^ but it was recently linked to negative regulation of the microbial response of alveolar macrophages. This feedback may however not function similarly in the host response to COVID-19 which is characterized by high influx of interstitial macrophages to the lung and may not be responsive to CC16 inhibition.

Overall, our comparative analysis suggests that while pathologically distinct entities, prolonged lung disease after acute COVID-19 may depend on the use of common features as IPF. Our study’s use of tissue from fatal cases of post-acute COVID-19 patients may not allow complete extrapolation to the full diversity of the PASC syndrome. It does however represent an extremely timely and valuable resource to study lung pathology of acute and post-acute COVID-19 from the microanatomical to single cell level and highlights persistent pathological derangement of lung structure in patients with prolonged lung disease post-acute COVID-19.

## Methods

### Data reporting

No statistical methods were used to predetermine sample size due to the limited availability of samples from post-acute COVID-19 patients.

### Tissue acquisition and preparation

Tissue samples were provided by the Weill Cornell Medicine Department of Pathology using protocols approved by the Tissue Procurement Facility of Weill Cornell Medicine. Experiments using samples from human subjects were conducted in accordance with local regulations and with the approval of the IRB at the Weill Cornell Medicine. The autopsy samples were collected under protocol 20-04021814.

Lung tissues were fixed via 10% neutral buffered formalin inflation, sectioned and fixed for 24 hours prior to processing and embedding into paraffin blocks. Freshly cut five micron sections were mounted onto charged slides.

### Antibody panel design

We designed an antibody panel capturing different immune and structural compartments of the lung. Antibody clones were validated through immunofluorescence and chromogenic staining and verified by a pathologist. 100 μg of purified antibody in BSA and Azide free format was conjugated using the MaxPar X8 multimetal labeling kit (Fluidigm) as per the manufacturer’s protocol. Sections were ablated on an Hyperion Imaging System (Fluidigm). To confirm the antibody binding specificity after conjugation and to identify the optimal dilution for each custom conjugated antibody, sections from the different patient tissues cohorts were stained. These sections were then ablated on Fluidigm Hyperion Imaging System and visualized using MCD Viewer for an expected staining pattern and optimal dilution that presented with good signal-to-noise ratio for each channel. For channels with visible spillover into the neighboring channels, a higher dilution factor was adopted when staining the cohort tissues.

### Imaging mass cytometry

Freshly cut 5-micron thick FFPE sections were stored at 4°C for a day before staining. Slides were first incubated for 1 hour at 60°C on a slide warmer followed by dewaxing in fresh CitriSolv (Decon Labs) twice for 10 minutes, rehydrated in descending series of 100%, 95%, 80%, and 75% ethanol for 5 minutes each. After 5 minutes of MilliQ water wash, the slides were treated with antigen retrieval solution (Tris-EDTA pH 9.2) for 30 minutes at 96°C. Slides were cooled to room temperature (RT), washed twice in TBS and blocked for 1.5 hours in SuperBlock Solution (ThermoFischer), followed by overnight incubation at 4°C with the prepared antibody cocktail containing all metal-labeled antibodies in a humid chamber (**Supplementary Table 2**). Next day, slides were washed twice in 0.2% Triton X-100 in PBS, and twice in TBS. DNA staining was performed using Intercalator-Iridium in PBS solution for 30 minutes in a humid chamber at room temperature. Slides were washed with MilliQ water and air dried prior to ablation. The instrument was calibrated using a tuning slide to optimize the sensitivity of detection range. Hematoxylin and Eosin (H&E) stained slides were used to guide the selection of regions representative of the whole range of lung pathology. All ablations were performed with a laser frequency of 200 Hz. Tuning was performed intermittently to ensure the signal detection integrity was within the detectable range.

### Immunohistochemistry

Automated immunohistochemistry on a Leica Bond III instrument was performed on five micron tissue sections using antibodies for myeloperoxidase (Clone 59A5, Leica ready to use antibody, without antigen retrieval) and CD163 (Clone MRQ-26, Leica ready to use antibody, antigen retrieval 20 minutes, high pH) using 3,3’ Diaminobenzidine chromogen. For each slide, a grid (5 × 5 grid, 0.4 cm × 0.4 cm boxes) was placed on the section and 5 alveolar, 2 vascular and 2 airway regions were randomly selected using random number generated X,Y coordinates and that region of interest (using a 20X objective) photographed.

### Preprocessing of imaging mass cytometry data

Imaging mass cytometry data was preprocessed as previously described^14^ with some modifications. Briefly, MCD files exported from the Hyperion instrument were converted to OME-TIFF format. To perform cell segmentation, we trained a pixel classifier using ilastik^56^ (version 1.3.3). We manually labeled pixels in random crops of images as belonging to one of three classes: nuclei – the area marked by signal in the DNA and Histone H3 channels; cytoplasm – the area immediately surrounding the nuclei and overlapping with signal in cytoplasmic channels; and background – pixels with low signal across all channels. The trained model was applied to the original images and output class probabilities for each pixel were then used to segment the images using DeepCell^57^ (version 0.12.1).

### Macroscopic assessment of imaging mass cytometry data

Calculation of the lacunar space in the images was done as previously described^14^. Briefly, we used the mean of all channels in each image stack after performing histogram equalization per channel and binarized the mean image with Otsu’s method. We then performed morphological dilation and closing operations with a disk of 5 μm diameter, filled holes in the image, and removed objects smaller than 625 μm (25 ** 2). The lacunar fraction of the image is the fraction of the image with positive signal.

To quantify fibrosis, we used the same definition as previously described^14^, except that we used both the Collagen Type I and Periostin channels. The final score is the average of the scores for both channels weighted by the average intensity of the channels across images.

### Annotation of microanatomical domains

To provide microanatomical context to the single-cells, we exported an JPEG image with the Aquaporin 1, Collagen type I, and alpha-smooth muscle actin channels, and used labelme^58^ to draw polygons representing specific areas of lung microanatomy as done before^59,60^. Polygons were converted into binary masks and intersected with the position of the cell segmentations for annotation of microanatomical domains per single-cell.

### Cell type identification

To identify cell types in an unsupervised fashion, we first quantified the intensity of each channel in each segmented cell not overlapping image borders. In addition, for each cell we computed various morphological features. Values were log-transformed, scaled per image with a cap at -3 and 3, and globally scaled. We used Combat^61^ to remove technical signal associated with specific samples, performed Principal Component Analysis (PCA), built a k-nearest neighbor graph using batch-balanced k-nearest neighbors (bbknn)^62^, performed Uniform Manifold Approximation and Projection (UMAP)^63^ (version 0.5.3), and finally clustered cells with the Leiden algorithm^64^ (*leidenalg* package, version 0.8.10) with resolution 3.5. All steps were performed with Scanpy^65^ (version 1.9.1). Each cluster was labeled with a cell type label based on the abundance of signal from the channels and the enrichment in each microanatomical domain. Only cell types with total abundance of at least 1% of total are displayed.

### Dimensionality reduction

We used various dimensionality reduction and manifold learning methods such as Multidimensional Scaling, Principal Component Analysis, Spectral Embedding, and Isomap from the scikit-learn package (version 1.0.2). The inputs were cell type abundance measurements for each region of interest where the abundance was normalized in percentage of total cells found per observation, and by area acquired. The two matrices were then concatenated, centered and scaled. Furthermore, in order to compare images with distinct microanatomical context, we regressed out the composition of each image based on the microanatomical annotations using the “regress_out” function in Scanpy. The same resulting matrix was also used for computation of pairwise correlations between images.

### Statistical testing

Unless otherwise mentioned, we used pairwise Mann-Whitney U-tests from the Pingouin package^66^ (version 0.5.2) to compare the abundance of cell types, positivity of markers, and abundance of interactions between groups of images. Reported effect size values are the Hedges g value. P-values were then adjusted for multiple testing with the Benjamini-Hochberg false discovery rate (FDR) method^67^. The results of testing are available in **Supplementary Table 3**.

## Data Availability

Multiplexed imaging data are available at the following publicly available repository: https://doi.org/10.5281/zenodo.7060271. Source code used in data analysis is available at the following publicly available repository: https://github.com/ElementoLab/post-covid-imc

https://github.com/ElementoLab/post-covid-imc

## Data availability

Multiplexed imaging data are available at the following publicly available repository: https://doi.org/10.5281/zenodo.7060271

## Code availability

Source code used in the analysis is available at the following publicly available repository: https://github.com/ElementoLab/post-covid-imc

## Acknowledgements

We thank the patients and their families for their contributions to our study. This work was partially supported by the WorldQuant Initiative for Quantitative Prediction. A.F.R. was supported by a NCI T32CA203702 grant. O.E. is supported by Volastra, Janssen and Eli Lilly research grants, NIH grants UL1TR002384 and R01CA194547, and Leukemia and Lymphoma Society SCOR 7012-16, SCOR 7021-20 and SCOR 180078-02 grants. R.E.S. is supported by NIH grants NIAID 2R01AI107301 and NIDDK R01DK121072 and the American Heart Association. R.E.S. is supported as Irma Hirschl Trust Research Award Scholar. We thank the Englander Institute of Precision Medicine Mass Cytometry Core facility for processing, labeling and acquiring samples for IMC imaging, and the translational research laboratory (Department of Pathology, Weill Cornell Medicine) for histology and IHC support.

## Supplementary Figures

**Supplementary Figure 1:**
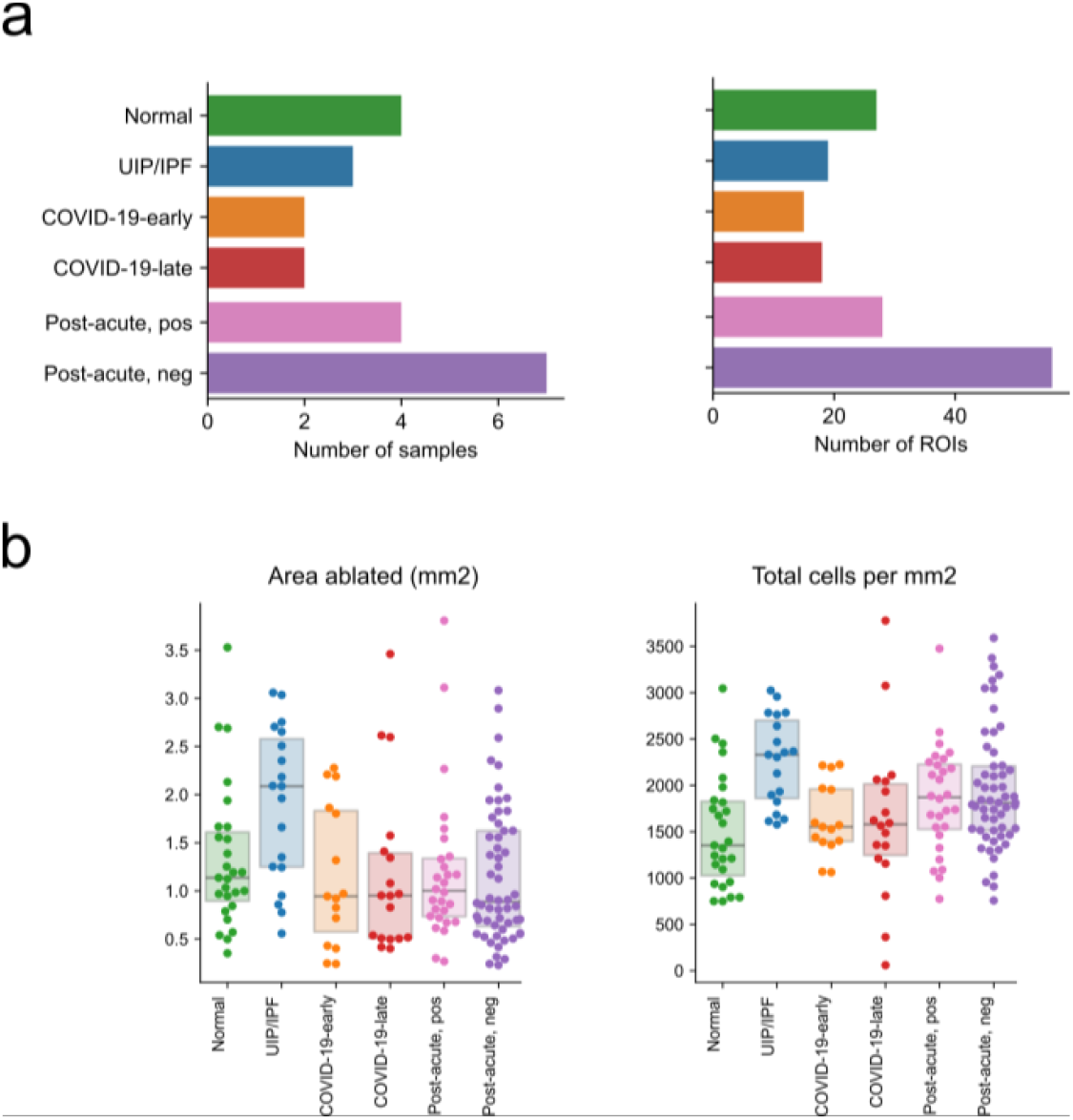
Technical properties of acquired multiplexed image data. **a**) Distribution of number of samples or regions of interest (ROI) per sample group. **b**) Image size (area ablated with imaging mass cytometry) (left), and cellular density (right) of images.

**Supplementary Figure 2:**
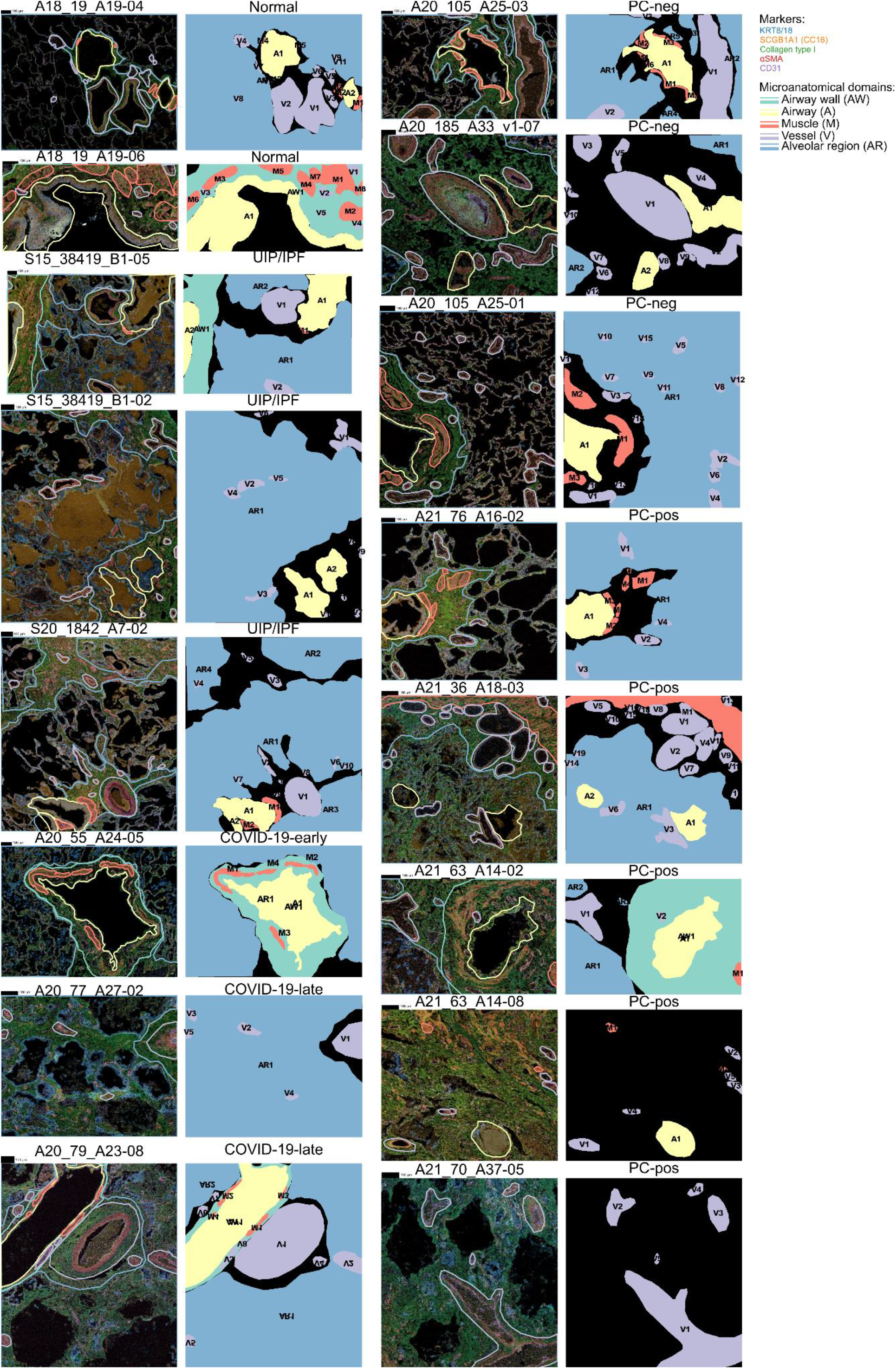
Microanatomical annotation of lung tissue. Images represent channel intensity overlaid with boundaries of microanatomical domains (left), and microanatomical domain areas (right). Areas in black represent unclear microanatomical context and cells under those areas were labeled with “remaining parenchyma” label.

**Supplementary Figure 3:**
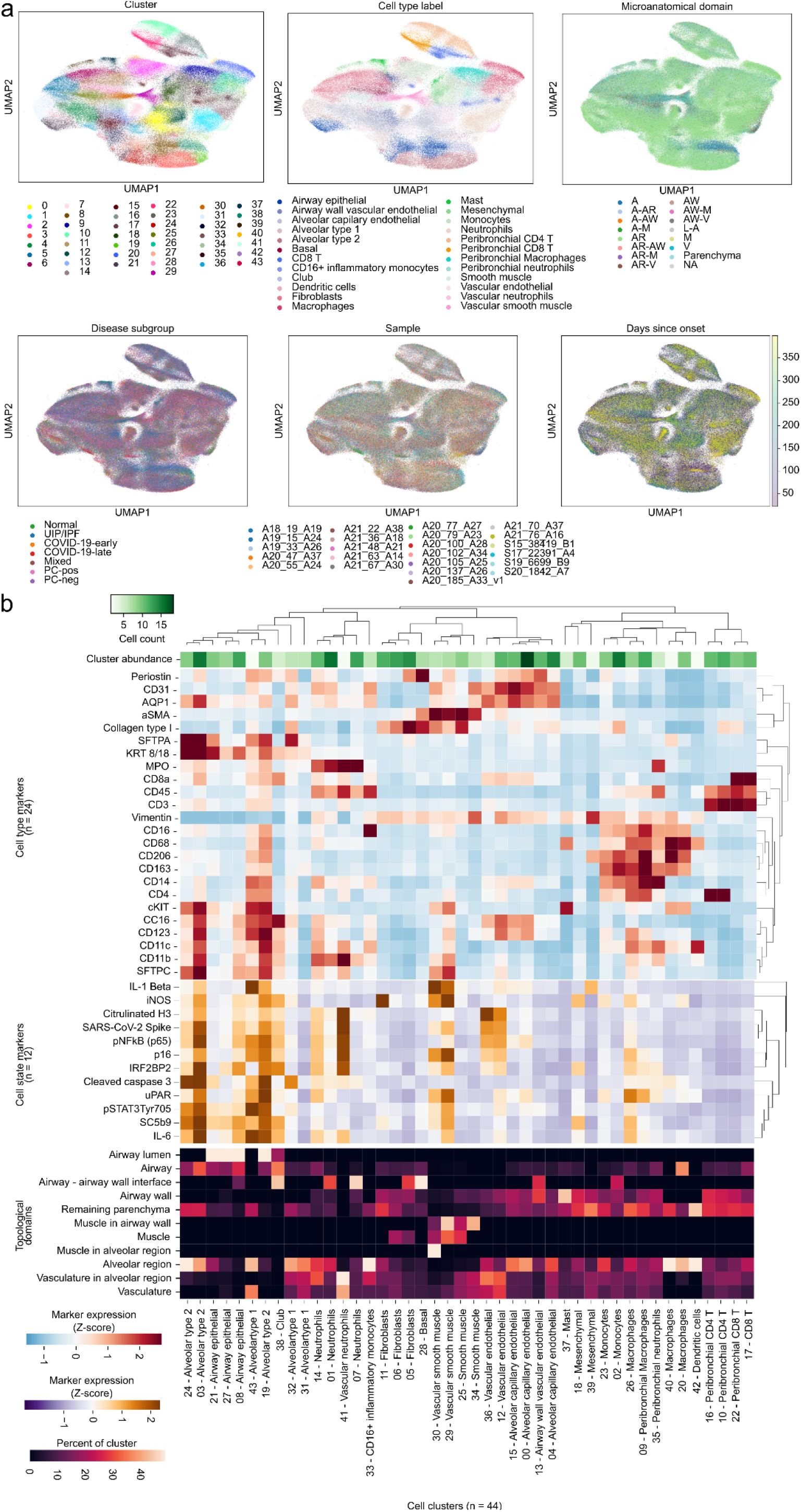
Phenotyping of single-cells detected with imaging mass cytometry. **a**) UMAP plot of single-cells colored by the clinical attributes of its sample, the microanatomical context of the cell, and by the clusters. **b**) Clustered heatmap of the discovered 44 clusters with the top part representing the average intensity of cell type markers in each cluster, the middle the average intensity of cell state markers, and the bottom the relative enrichment in microanatomical structures for each cell cluster.

**Supplementary Figure 4:**
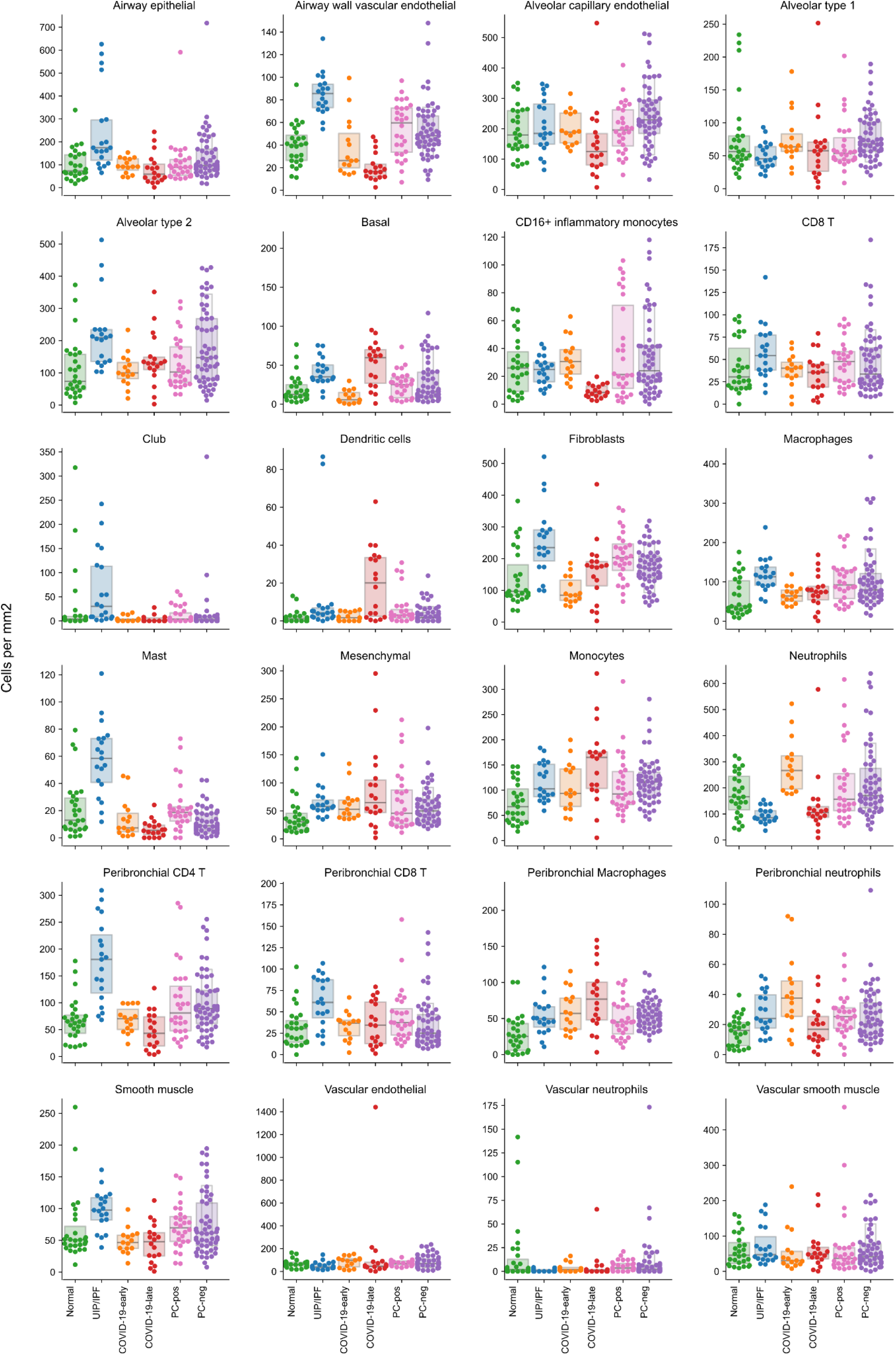
Changes in cell type abundance between sample groups. Cell density (cells per square millimeter) of each cell type for each sample group.

**Supplementary Figure 5:**
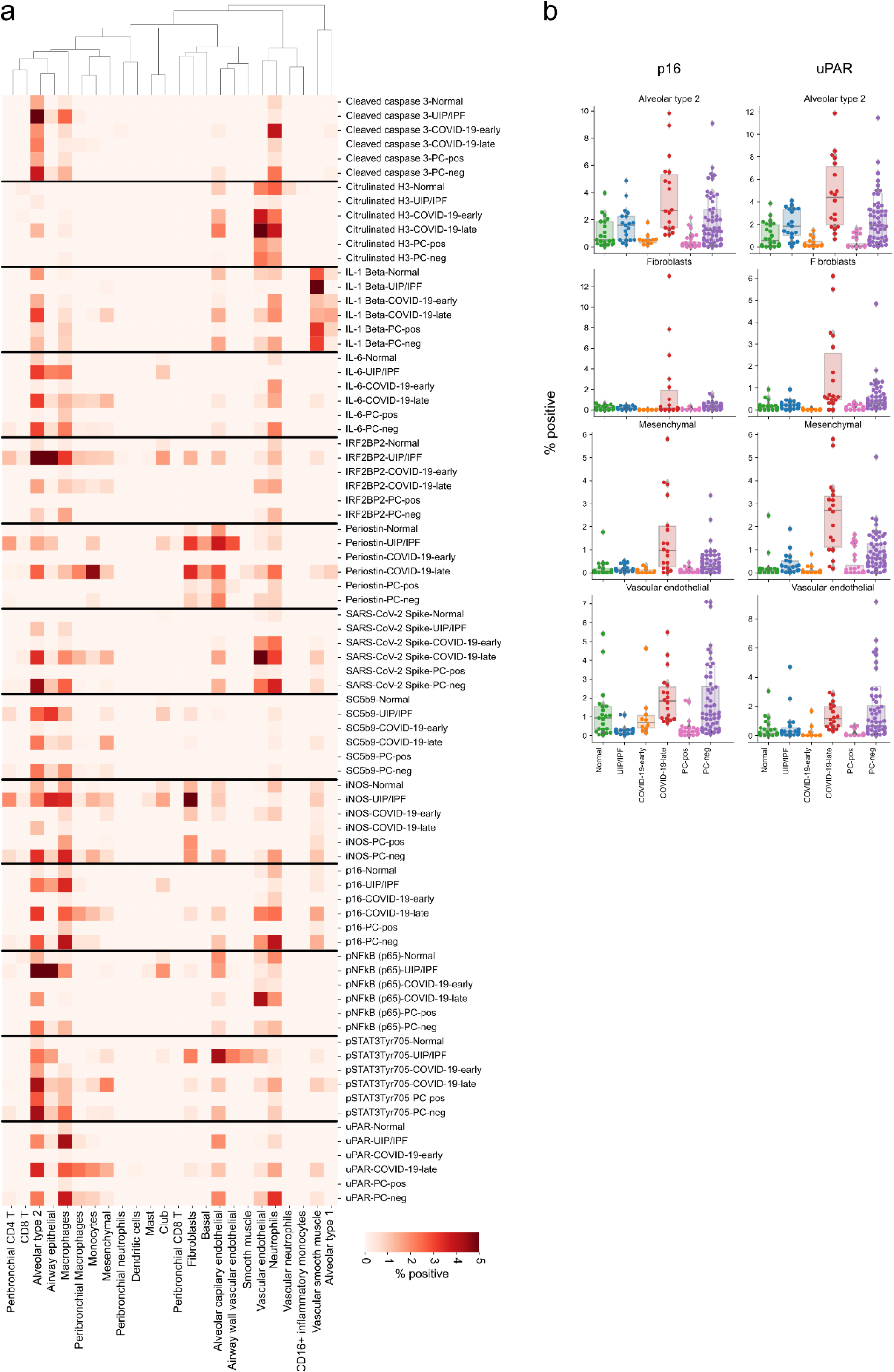
Changes in expression of cell state markers between sample groups. Percentage of cells positive for a marker for each cell type in each sample group.

**Supplementary Figure 6:**
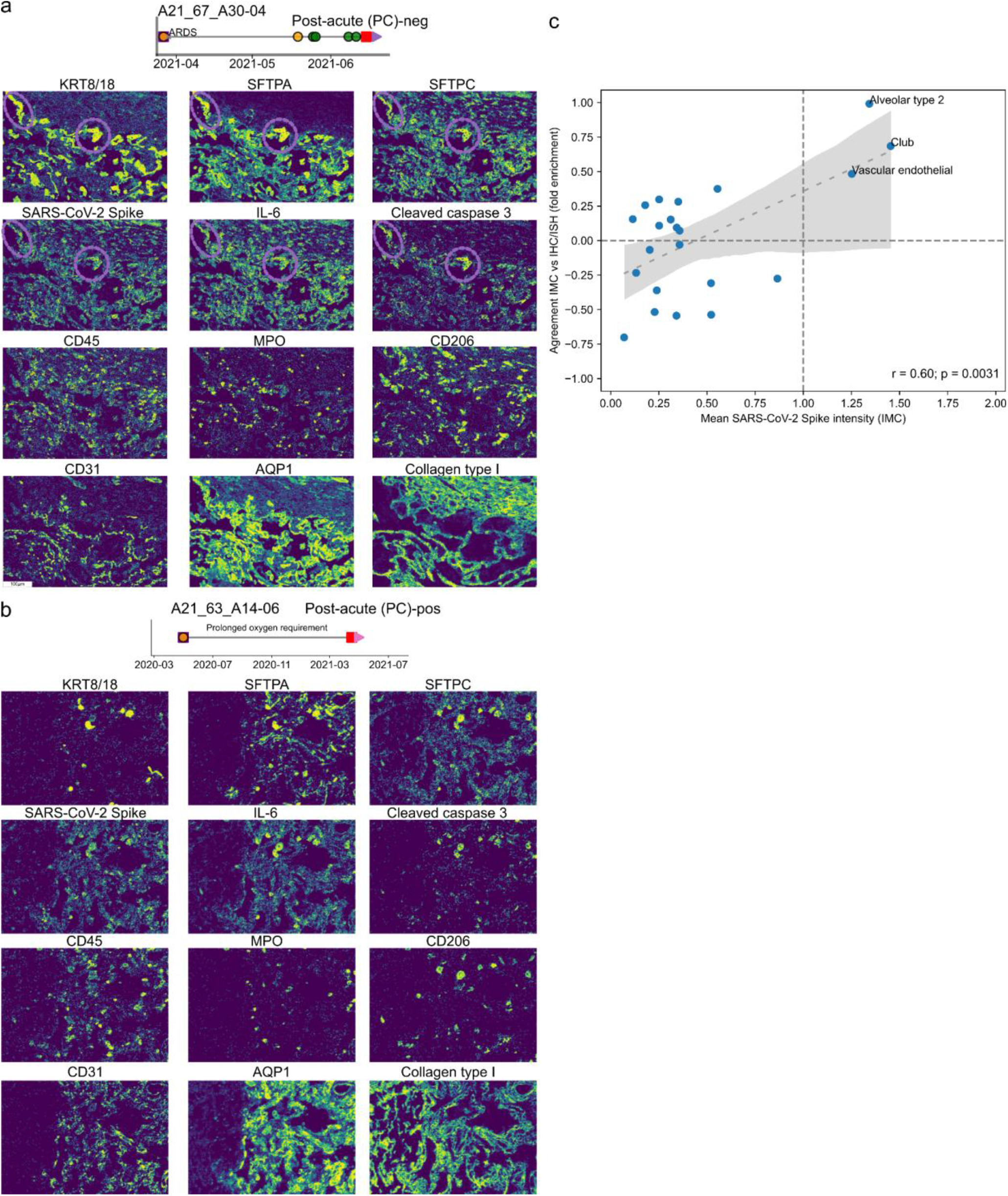
Detection and validation of SARS-CoV-2 proteins in tissue. **a-b**) Selected images illustrating SARS-CoV-2 signal in lung tissue from a PC-neg (a) and PC-pos (b) patient. **c**) Agreement between orthogonal methods and multiplexed imaging in quantification of SARS-CoV-2 detection.

**Supplementary Figure 7:**
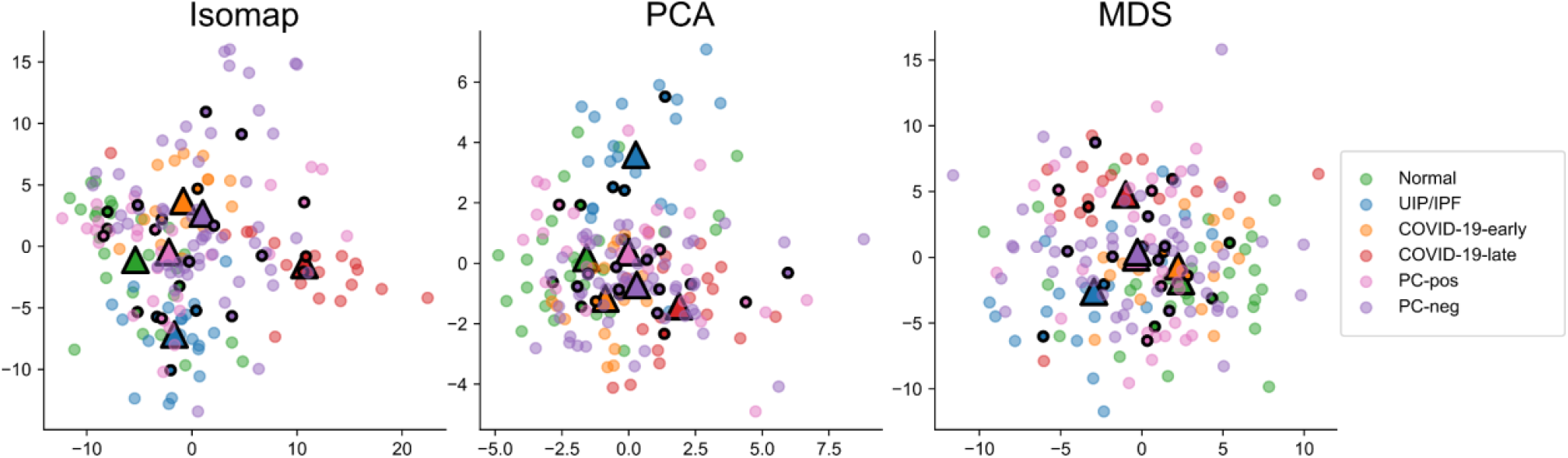
Global similarity between sample groups given by various dimensionality reduction methods. For Principal Component Analysis only the first two dimensions are shown.

## Supplementary Tables

**Supplementary Table 1.** Demographic and clinical details of samples included in analysis.

| ID | Group | Fever | Cough | Shortness of breath | Days intubated | Days of disease | Days in hospital | Days since onset to death | PLT/ mL | D-dimer (mg/L) | WBC | LY % | PMN % | CRP mg/L | ESR mm/h |
| --- | --- | --- | --- | --- | --- | --- | --- | --- | --- | --- | --- | --- | --- | --- | --- |
| 3 | COVID-19-early | Y | Y | Y | 0 | 5 | 0 |  | NA | NA | NA | NA | NA | NA | NA |
| 10 | COVID-19-early | Y | Y | Y | 0 | 11 | 5 |  | 400 | 33122 | 9.5 | 23 | 68 | 24.6 | NA |
| 28 | COVID-19-late | Y | N | Y | 32 | 36 | 36 |  | 348 | 4848 | 15.8 | 0.9 | 97.1 |  |  |
| 30 | COVID-19-late | 39 | Y | Y | 29 | 35 | 35 |  | 126 | 5128 | 11.5 | 0.5 | 91.7 |  |  |
| 64 | PC |  |  |  | 30 | 30 | 30 | 64 | 213 | 270 | 5.66 | 16 | 78.5 | 16.07 | 71 |
| 38 | PC-neg | 38.2 | N | Y |  | 44 |  | 44 | 212 | 754 | 8.3 | 17 | 74.3 | 1.3 | NA |
| 39 | PC-neg |  | Y | Y | post covid | post covid | post covid | 81 | 271 | 3996 | 9.3 | 3.9 | 86.5 | 4.1 |  |
| 40 | PC-neg | N | N | N | post covid | post covid | post covid | 29 |  |  |  |  |  |  |  |
| 5 | PC-neg | 36 | N | N | 28 | post covid | 28 | 120 | 304 | 1860 | 35.7 | 43 | 42 | NA | NA |
| 55 | PC-neg |  |  |  | NA | NA | NA | 200 | NA | NA | NA | NA | NA | NA | NA |
| 51 | PC-neg | Y | Y | Y | 30 | 50 | 43 | 42 | 300 | 3816 | 16.3 | 5.5 | 90.1 | 15.6 | na |
| 69 | PC-neg | Y | N | N | 0 | 5 | 22 | 22 | 269 | 2307 | 9.8 | 6.7 | 83 |  | 66 |
| 68 | PC-pos | Y | Y | N | 0 | NA | 0 | 350 | 361 | 1067 | 3.8 | 12 | 74.6 | 15 | 121 |
| 61 | PC-pos | Y | Y | Y | NA | NA | NA | 350 |  |  |  |  |  |  |  |
| 70 | PC-pos | N | Y | Y | 22 | 40 | 30 | 155 | 150 | 18538 | 6.4 | 12 | 82 | 5.27 | 72 |
| 71 | PC-pos | N | N | Y | 0 | 18 | 18 | 399 | 165 | 5460 | 8.28 | 28 | 58 | 7 | 100 |

**Supplementary Table 2.**
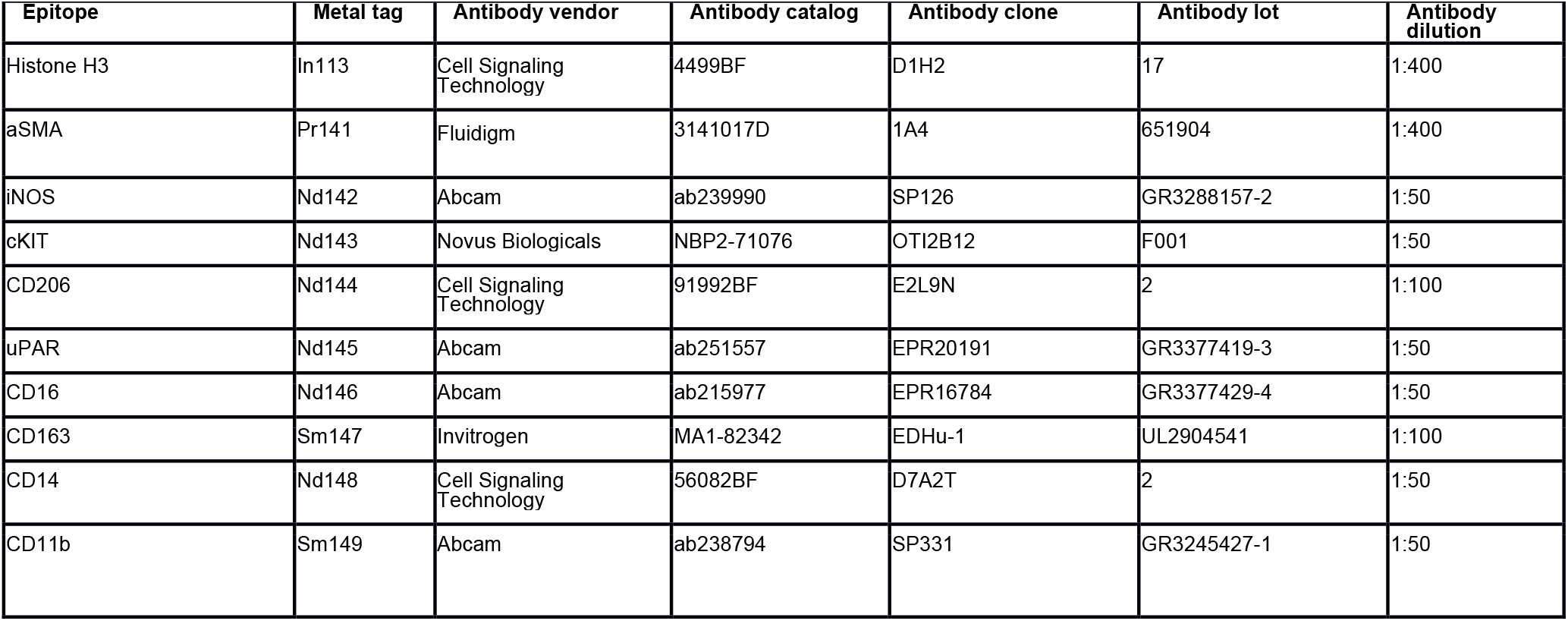
Markers used in imaging mass cytometry analysis of lung tissue.

**Supplementary Table 3.**
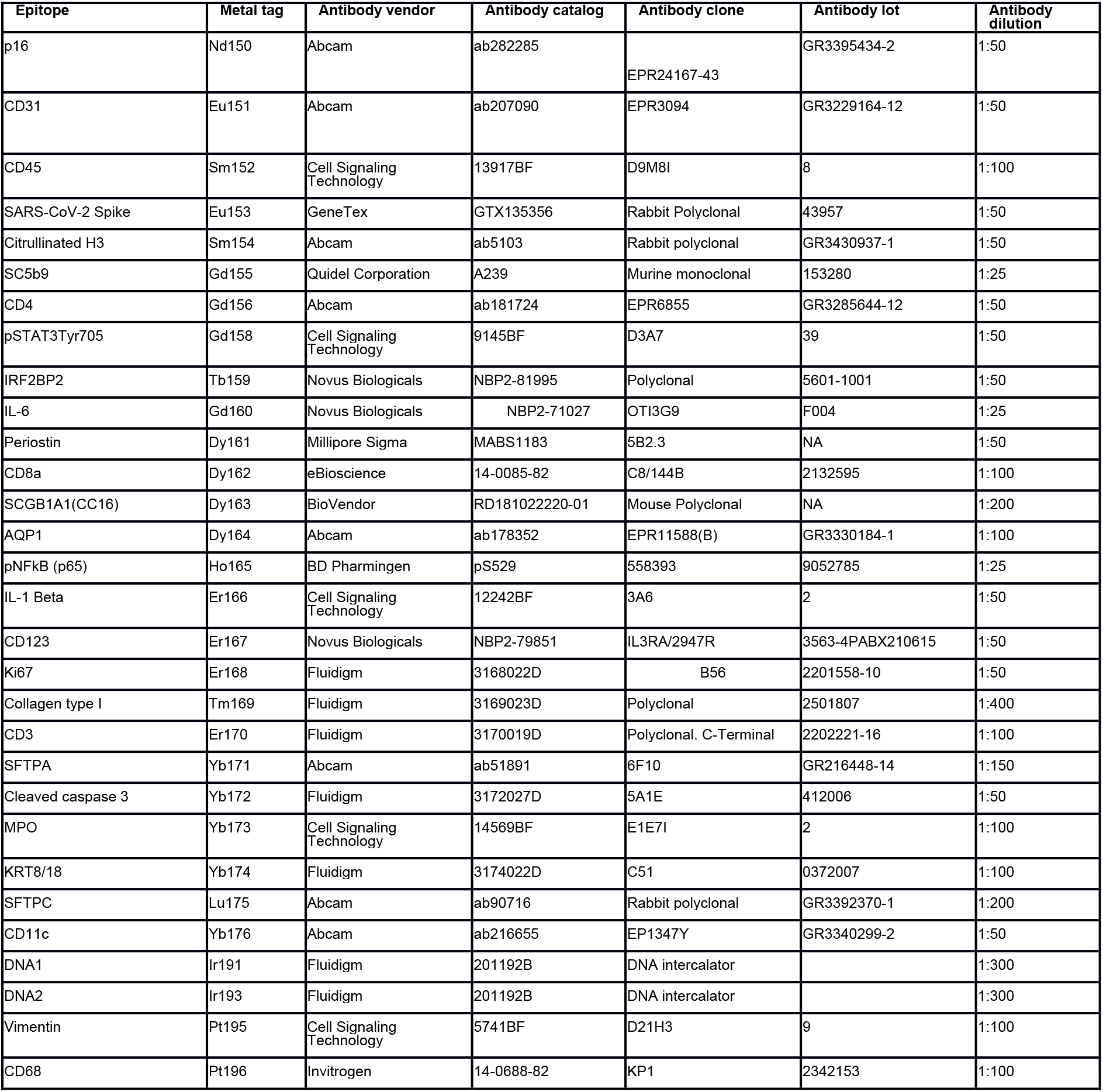
Statistical testing for differences between sample groups. Testing was performed for differences in lacunarity, fibrosis, cellular abundance and marker positivity. For cell type abundance comparisons values tested are absolute frequencies (cells per square millimeter). The table is available as an external file to the manuscript.

