## Supplementary figures and images for "Persistent alveolar type 2 dysfunction and lung structural derangement in post-acute COVID-19"

### Supplementary Figure 1

#### Supplementary Figure 01

a

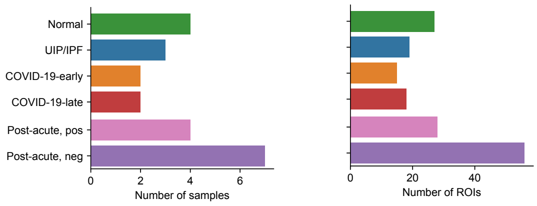


b

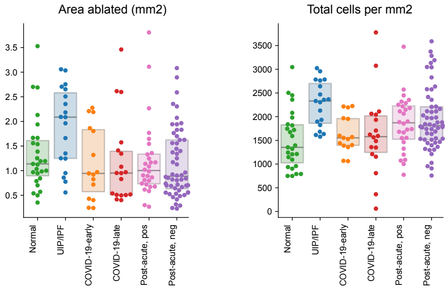

### Supplementary Figure 2

#### Supplementary Figure 02

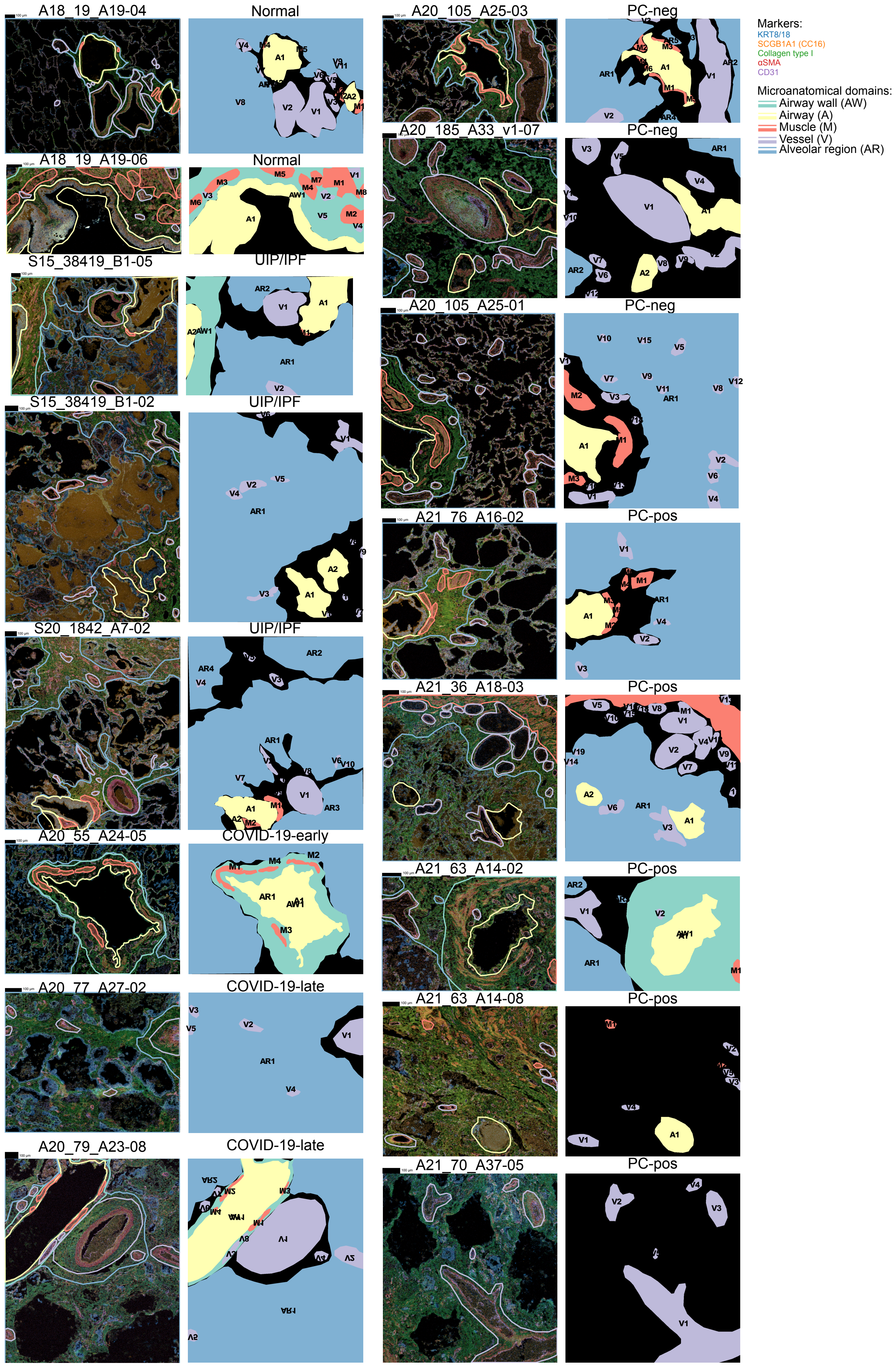

### Supplementary Figure 3

#### Supplementary Figure 03

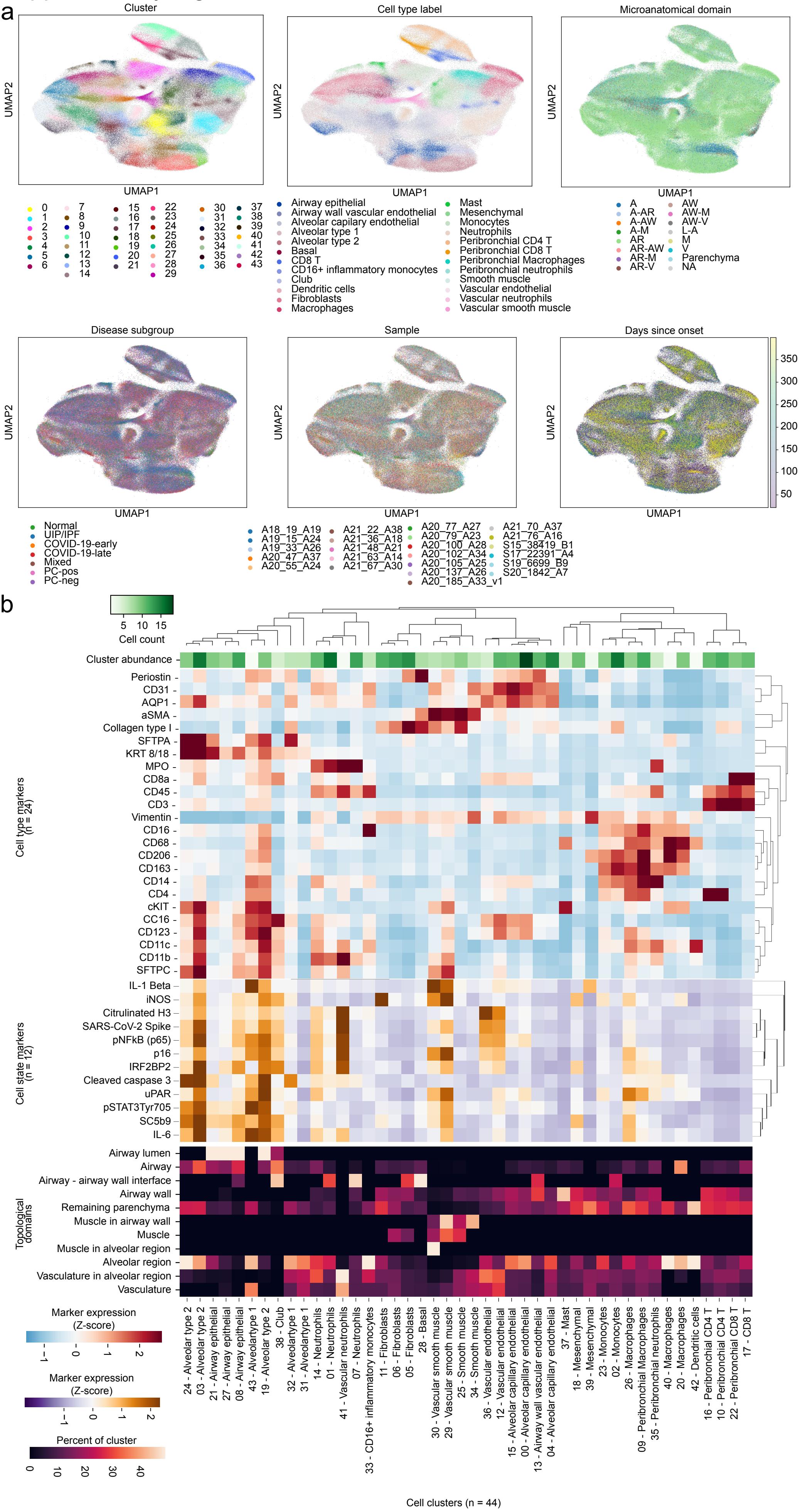

### Supplementary Figure 4

#### Supplementary Figure 04

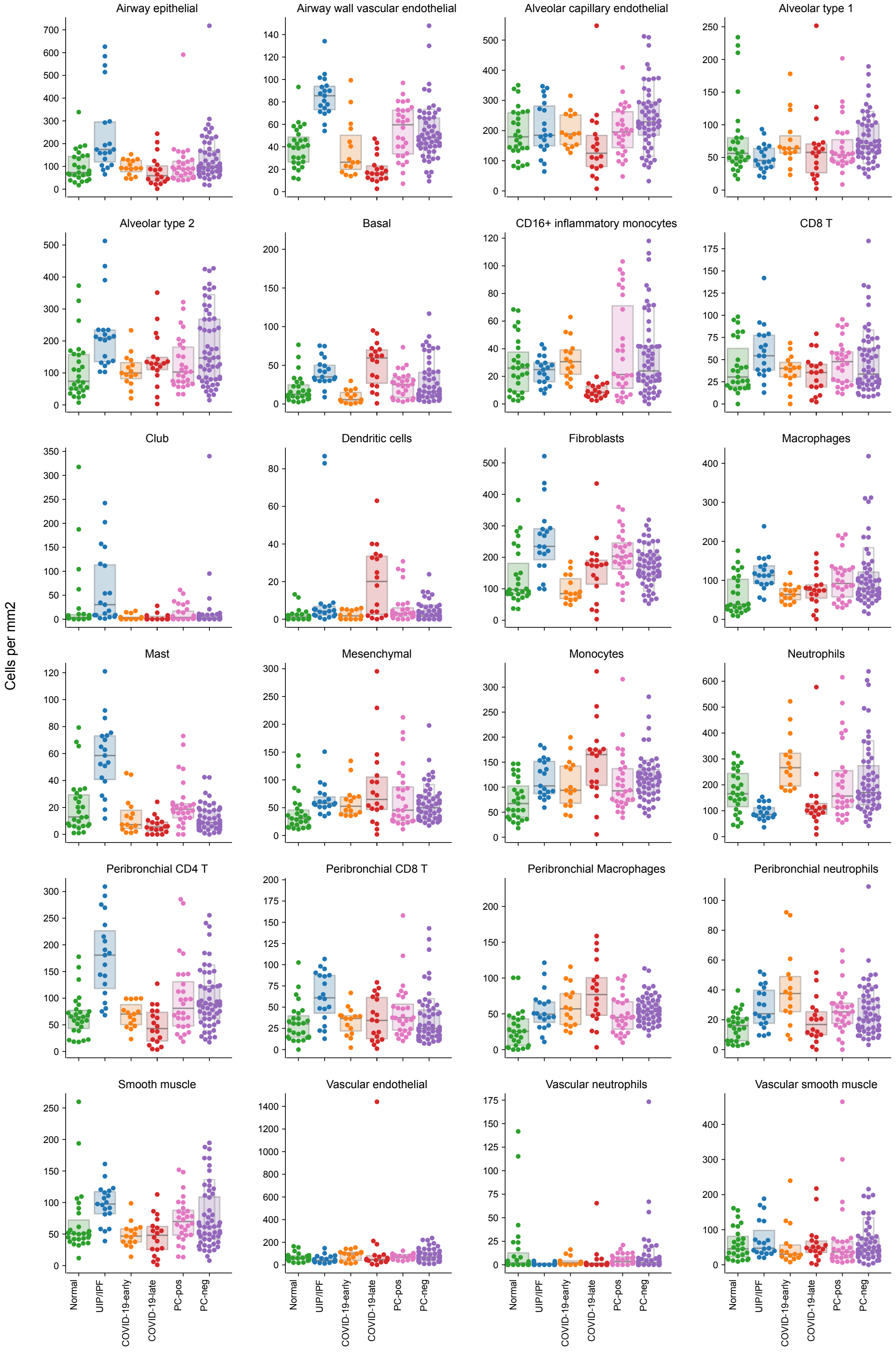

### Supplementary Figure 5

#### Supplementary Figure 05

a

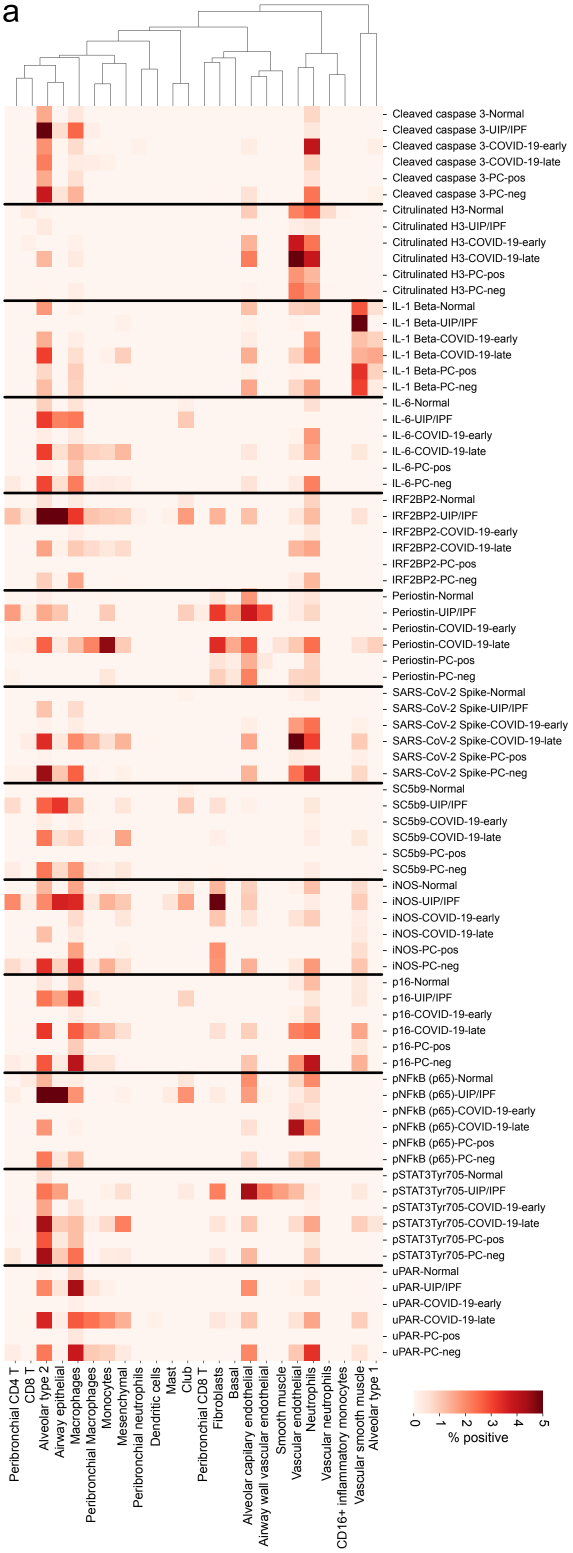


b

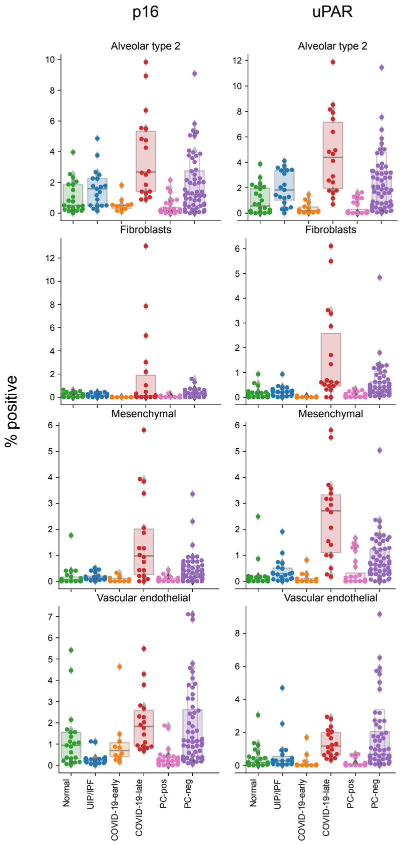

### Supplementary Figure 6

Supplementary Figure 06

a

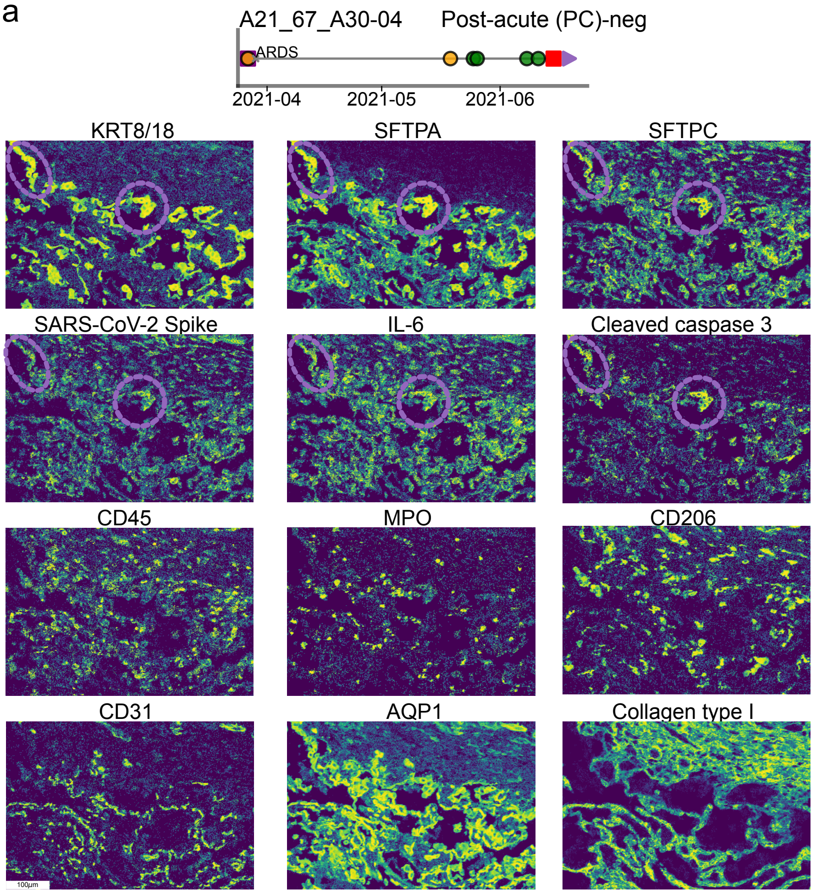


b

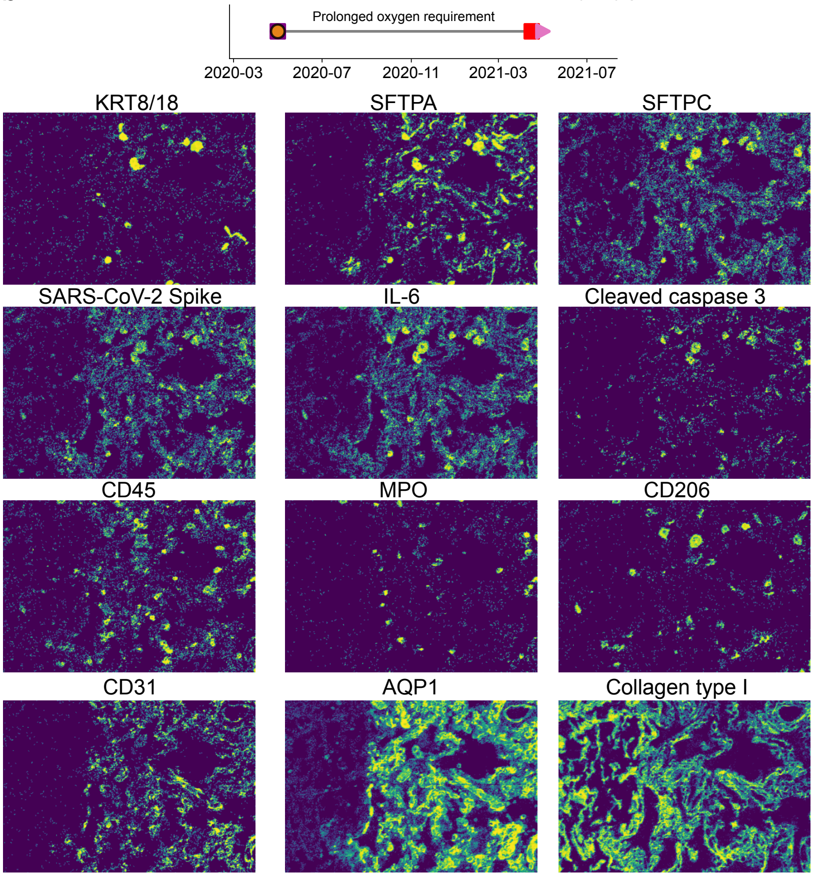


c

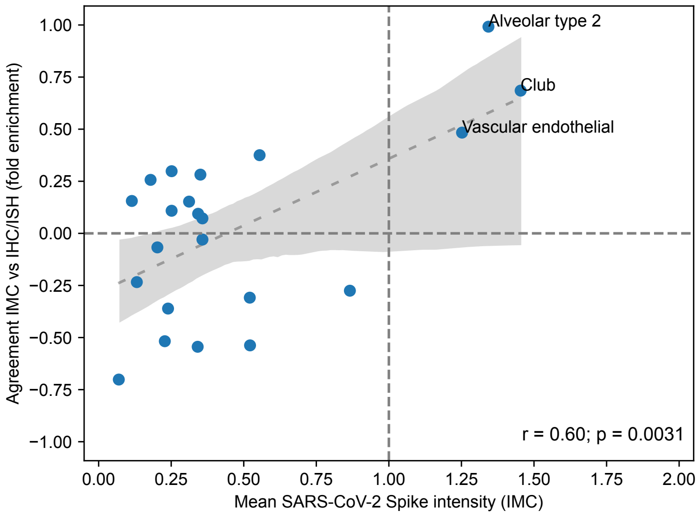

### Supplementary Figure 7

Supplementary Figure 07

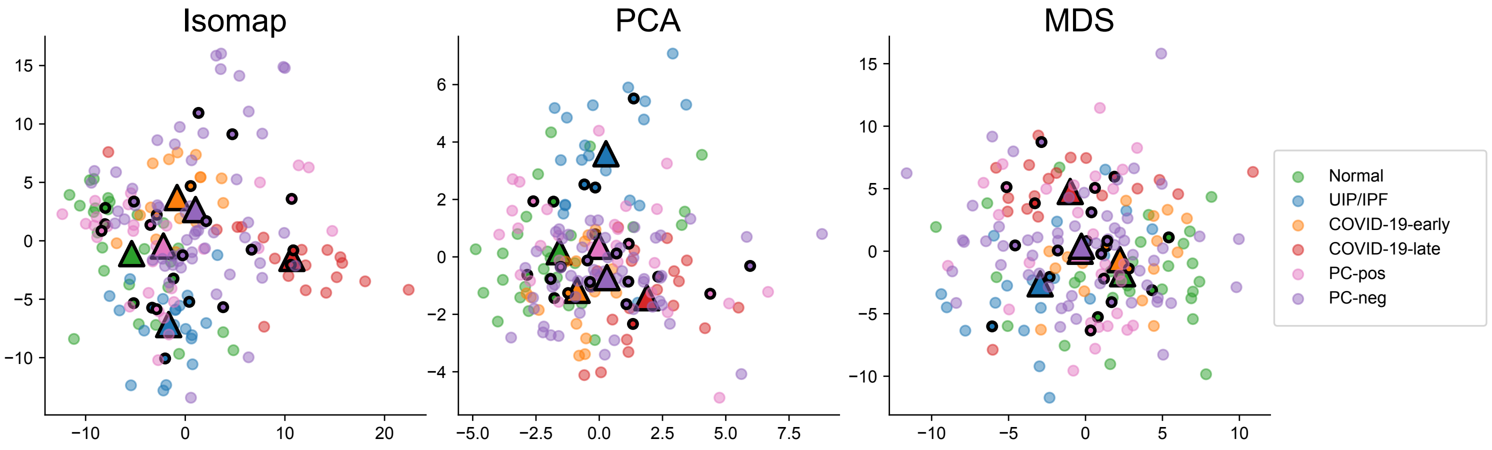
